# Delirium is a presenting symptom of COVID-19 in frail, older adults: a cohort study of 322 hospitalised and 535 community-based older adults

**DOI:** 10.1101/2020.06.15.20131722

**Authors:** Maria Beatrice Zazzara, Rose S. Penfold, Amy L. Roberts, Karla A. Lee, Hannah Dooley, Carole H. Sudre, Carly Welch, Ruth C. E. Bowyer, Alessia Visconti, Massimo Mangino, Maxim B. Freydin, Julia S. El-Sayed Moustafa, Kerrin Small, Benjamin Murray, Marc Modat, Jonathan Wolf, Sebastien Ourselin, Finbarr C. Martin, Claire J. Steves, Mary Ni Lochlainn

## Abstract

**Background:** Frailty, increased vulnerability to physiological stressors, is associated with adverse outcomes. COVID-19 exhibits a more severe disease course in older, co-morbid adults. Awareness of atypical presentations is critical to facilitate early identification.

**Objective:** To assess how frailty affects presenting COVID-19 symptoms in older adults.

**Design:** Observational cohort study of hospitalised older patients and self-report data for community-based older adults.

**Setting:** Admissions to St Thomas’ Hospital, London with laboratory-confirmed COVID-19. Community-based data for 535 older adults using the COVID Symptom Study mobile application.

**Subjects:** <u>Hospital cohort:</u> patients aged 65 and over (n=322); unscheduled hospital admission between March 1^st^, 2020-May 5^th^, 2020; COVID-19 confirmed by RT-PCR of nasopharyngeal swab. <u>Community-based cohort:</u> participants aged 65 and over enrolled in the COVID Symptom Study (n=535); reported test-positive for COVID-19 from March 24th (application launch)-May 8^th^, 2020.

**Methods:** Multivariate logistic regression analysis performed on age-matched samples from hospital and community-based cohorts to ascertain association of frailty with symptoms of confirmed COVID-19.

**Results:** <u>Hospital cohort:</u> significantly higher prevalence of delirium in the frail sample, with no difference in fever or cough. <u>Community-based cohort</u> :significantly higher prevalence of probable delirium in frailer, older adults, and fatigue and shortness of breath.

**Conclusions:** This is the first study demonstrating higher prevalence of delirium as a COVID-19 symptom in older adults with frailty compared to other older adults. This emphasises need for systematic frailty assessment and screening for delirium in acutely ill older patients in hospital and community settings. Clinicians should suspect COVID-19 in frail adults with delirium.

## Background

COVID-19, an acute respiratory disease caused by severe acute respiratory syndrome coronavirus 2 (SARS-CoV-2), poses a significant threat to the older population, who are at increased risk from infection, hospitalisation and death[1,2]. In a study from Wuhan, China, over 70% of 339 patients over 60 years of age with COVID-19 were classified as severe or critical, with a case fatality rate of 19% and median survival of 5 days following admission[3]. In Italy, the highest case fatality rate was observed among older, male patients with multiple comorbidities[4] and in an American study, adults >65 years old represented 80% of COVID-19 fatalities[5]. Vulnerability within the older population might partially be explained by a greater number and severity of chronic diseases[6] and immunosenescence[7,8]. It is imperative to rapidly identify and contain infection within this group.

Prevalent symptoms reported in patients with confirmed COVID-19 infection from a variety of international settings are summarised in Table 1. However, it is well-documented that presenting symptoms vary with age for many diseases, including respiratory infections[9,10,11]. Symptoms including delirium, generalised weakness, malaise, anorexia, headache, dizziness, falls and functional decline are more common in older people. Less typical presentations of COVID-19 in older adults have been documented in case reports and small cohort studies (n<60), with symptoms including weakness, headache, delirium and an absence of fever or cough[12,13,14]. In work by our research group using point-of-care testing for COVID-19 in older adults, hypothermia was noted to be an early clinical sign in a minority of patients[15].

**Table 1.** Studies describing the frequency of symptoms in laboratory confirmed SARS-CoV-2

| **Study** | **Participants** | **Study Methods** | **Symptoms reported (% of cases)** |
| --- | --- | --- | --- |
| Guan et al.[26]<br>China | 1099 patients with laboratory confirmed SARS-CoV-2<br>Median age 47 years<br>58.1% Male<br>Severe disease<br>n = 173<br>Non-severe disease<br>n = 926 | 552 hospitals<br>Retrospective observational study<br>Data compared between severe and non-severe disease (severe = death or requirement for ICU admission/mechanical ventilation) | Fever (43.8% on admission, 88.7% during hospitalization)<br>Cough (67.8%)<br>Fatigue (38.1%)<br>Sputum production (33.7%)<br>Shortness of breath (18.7%)<br>Myalgia or arthralgia (14.9%)<br>Sore throat (13.9%)<br>Headache (13.6%)<br>Chills (11.5%)<br>Nausea or vomiting (5.0%)<br>Diarrhea (3.8%) |
| Wang et al. [49]<br>China | 138 patients<br>Median age 56 years<br>54.3% Male | Single site<br>Retrospective observational study | Fever (98.6%)<br>Cough (86.2%)<br>Myalgia or fatigue (34.8%)<br>Dyspnea (31.2%)<br>Expectoration (26.8%)<br>Sore throat (17.4%)<br>Diarrhea (10.1%)<br>Headache or dizziness (6.5%)<br>Nausea and vomiting (3.6%)<br>Other symptoms* (42.0%)<br>* chilliness; conjunctival congestion; anorexia; abdominal pain; constipation; heart palpitations |
| Du et al.[50]<br>China | 85 patients with laboratory confirmed SARS-CoV-2<br>Median age 65.8 years<br>72.9% Male | 2 hospitals<br>Retrospective observational study<br>All cases fatal | Fever (91.8%)<br>Dyspnea (70.6%)<br>Shortness of breath (58.8%)<br>Fatigue (58.8%)<br>Anorexia (56.5%)<br>Expectoration (37.6%)<br>Dry cough (22.4%)<br>Diarrhea (18.8%)<br>Myalgia (16.5%)<br>Headache (4.7%)<br>Vomiting (4.7%)<br>Abdominal pain (3.5%)<br>Chest pain (2.4%)<br>Pharyngalgia (2.4%) |
| Liu et al. [51]<br>China | 32 patients with laboratory confirmed SARS-CoV-2 (25 adults)<br>Mean age 44 years<br>48% Male | Single site<br>Retrospective observational study | Fever (94%)<br>Cough (76%)<br>Myalgia or fatigue (52%)<br>Shortness of breath (40%)<br>Pharyngalgia (24%)<br>Diarrhea or vomiting (8%) |
| Huang et al. [52]<br>China | 32 patients with laboratory confirmed SARS-CoV-2<br>Median age 49 years<br>73% Male | Single site<br>Retrospective and prospective observational study<br>Data compared between cases requiring ICU care and no ICU care | Fever (98%)<br>Cough (76%)<br>Myalgia or fatigue (44%)<br>Sputum production (28%)<br>Headache (8%)<br>Hemoptysis (5%)<br>Diarrhea (3%) |
| Xu et al. [53]<br>China | 62 patients with laboratory confirmed SARS-CoV-2<br>Median age 41 years<br>56% Male | Seven hospitals<br>Retrospective case series | Cough (81%)<br>Fever (77%)<br>Expectoration (56%)<br>Myalgia or fatigue (52%)<br>Headache (34%)<br>Diarrhea (8%)<br>Hemoptysis (3%)<br>Shortness of breath (3%) |
| SARS-CoV-2 Surveillance Group Istituto Superiore di Sanità[54] (23<sup>rd</sup> April 2020)<br>Italy | 23,188 patients with PCR-confirmed SARS-CoV-2<br>Median age 81 years<br>63.3% Male | Patients dying in Italy: 19 regions and 2 autonomous provinces<br>Retrospective data collection | Fever (75%)<br>Dyspnea (72%)<br>Cough (38%)<br>Diarrhea (6%)<br>Hemoptysis (1%) |
| Spiteri et al. [55]<br>WHO European Region | 38 cases with laboratory-confirmed SARS-CoV-2 (31 with recorded symptoms)<br>Median age 42 years<br>65.8% Male | Multiple centers<br>Retrospective observational study | Fever (64.5%)<br>Cough (45.2%)<br>Weakness (25.8%)<br>Headaches (19.4%)<br>Sore throat (6.5%)<br>Rhinorrhea (6.5%)<br>Shortness of breath (6.5%)<br>Asymptomatic (6.5%) |
| Zayet et al.[56]<br>France | 62 patients<br>Median age 56 years<br>39% Male | Single site<br>Retrospective observational study | Fatigue (94%)<br>Cough (81%)<br>Headache (78%)<br>Fever (76%)<br>Myalgia (61%)<br>Anosmia (52%)<br>Dysgeusia (48%)<br>Rhinorrhea (48%)<br>Diarrhea (39%)<br>Dyspnea (34%) |

Frailty, an increased vulnerability to physiological stressors[16], confers increased risk of adverse health outcomes including mortality, falls, fractures, comorbidities, hospitalisation, and disability[16,17]. Frailty is also associated with increased likelihood of hospital-acquired complications, such as delirium, falls or deconditioning. Recent National Institute for Health and Care Excellence (NICE) guidelines in the UK recommend that all adult patients over 65 years old should be assessed for frailty on hospital admission to guide appropriate management and advanced care planning, using the Clinical Frailty Scale (CFS) where relevant[18]. However, no study has reported on effects of frailty on symptoms of COVID-19, despite calls for data in this important area[19].

We use point-of-care data from patients admitted to a large UK hospital trust, supported by community-based COVID Symptom Study mobile application (“app”) data, to assess how frailty affects presentation of confirmed COVID-19 infection in older adults.

## Methods

### Participants

This study uses two cohorts.

#### (i) Hospital cohort

Point-of-care clinical data was collected on all patients aged 65 years and above with unscheduled admission to Guy’s and St Thomas’ Hospital, London, between March 1^st^-May 5^th^, 2020 with suspected or confirmed COVID-19 infection. This was collected as part of the international COVIDCollab study, led by the Geriatric Medicine Research Collaborative[20]. All data is routinely collected and includes age, sex, body mass index (BMI), co-morbidities, CFS[21,22] and presenting symptoms. Full data collection template available at https://www.gemresearchuk.com/covid-19.

Only laboratory-confirmed cases of COVID-19 infection (RT-PCR of nasopharyngeal swab) were included in this study. For patients readmitted during the study period, data from index admission was used.

#### (ii) Community-based cohort

This comprised community-dwelling older people who had submitted data via the COVID Symptom Study app, developed by Zoe Global Limited, with scientific input from researchers and clinicians at King’s College London and Massachusetts General Hospital, Boston. The app was launched in the UK on March 24^th^ and became available in the US on March 29^th^, 2020. It captures self-reported information on COVID-19 symptoms and healthcare visits. On first accessing the app, users record location, age and health risk factors. Since the CFS requires assessment by a healthcare professional, it was not feasible here. Users instead completed a PRISMA-7 questionnaire (a 7-item screening tool)[23] adapted for large-scale app usage, to assess level of frailty (Supplementary Table SP1). PRISMA-7 has been validated for use in older adults[24] and has good validity compared to the CFS[25]. The app was upgraded in April 2020 to allow family members or care-givers to submit data by-proxy, with consent. This has increased numbers of reporting older, frailer participants.

Consent to data analysis and sharing was obtained when individuals enrolled in the app. UK data up to May 8^th^, 2020 were inspected and, as part of the quality control (QC) stage, invalid BMI (<15 and >55) and age entries (<18 years) removed. QC was performed by an automated python script developed independent of data analysis. The total dataset comprised 2,848,396 individuals, 11,206 of whom had reported a positive COVID-19 test. This was filtered to include only those aged 65 years and above. All longitudinal entries were condensed per person to form a binary metric recording whether the person had experienced each of the 14 symptoms (below) during the study period.

### Measurements

Delirium was defined as a clinically documented delirium diagnosis on admission in the hospitalised cohort and “Yes” to the following question in the community-based cohort: ‘Do you have any of the following symptoms: confusion, disorientation or drowsiness?’

### Analyses

Descriptive statistics were used to describe demographic and key clinical characteristics of the study populations: overall and according to frailty. Frailty was classified as a binary variable: CFS≥ 5 = frail; CFS<5 = not frail for the hospital cohort and PRISMA7≥3 = frail; PRISMA7<3 = not frail in the community-based cohort. A binary temperature classification was used in the hospitalised cohort, based on a previous large observational cohort study[26]: ≥37.5°C = fever; 37.5 < afebrile.

Continuous variables were expressed as mean +/- standard deviation, and categorical variables as frequencies by absolute value and percentages (%). Differences in proportions and means of covariates between subjects with and without frailty were assessed using Wilcoxon’s test for continuous variables and Fisher’s exact test for categorical variables.

Following age-matching by frailty status in the hospitalised cohort (n=210), multivariable logistic regression analysis was performed to assess whether frailty predicts symptoms of confirmed COVID-19 infection on hospital admission, including fever, cough and delirium. BMI and sex were covariates.

Following age-matching by frailty status in the community-based cohort (n=238), multivariable logistic regression analysis was performed to assess whether frailty predicts symptoms of reported confirmed COVID-19 infection. Symptoms analysed included delirium, fever, persistent cough, fatigue, myalgia, shortness of breath, diarrhea, anorexia, abdominal pain, hoarse voice, anosmia/dysgeusia, headache, chest pain and pharyngodynia. BMI and sex were covariates.

Data analysis and graphics were performed in the R statistical environment (version 4.0) using the Tidyverse packages.

## Results

### Hospital cohort

Baseline characteristics of the unmatched hospital study cohort are summarised in Supplementary Table SP2. Mean age of 322 individuals with confirmed COVID-19 RT-PCR was 78.58 years (standard deviation (SD) +/- 7.93); 154 (48%) were females. 165 (51%) patients were classified as frail.

After age-matching, difference in mean age between the frail (77.92 +/- SD 6.64 years) and non-frail group (77.85 +/- SD 7.04 years) was non-significant (Wilcoxon’s *p*>0.05). The proportion of females was 39% in both groups. BMI differences between the frail and non-frail groups were non-significant (Figure 1 a-c and Table 2). Comorbidities of the matched study cohort according to frailty status are summarised in Table 2. There was a higher proportion of respiratory disease and dementia in the frail compared to non-frail group, but no significant differences in prevalence of cardiovascular disease, diabetes, psychiatric disease or cancer (Table 2).

**Table 2:** Characteristics and comorbidities of age-matched hospital and community-based cohorts, according to frailty status. Binary variables are presented as count (%) and continuous variables as mean (standard deviation). Frail Hospital = CFS ≥ 5; Frail Community= PRISMA7 ≥3.

|  | Hospital Cohort |  |  |  | Community Cohort |  |  |  |
| --- | --- | --- | --- | --- | --- | --- | --- | --- |
|  | Total Sample<br>(n = 210) | Frail<br>(n = 105) | Not Frail<br>(n = 105) | P-value | Total Sample<br>(n=238) | Frail<br>(n=119) | Not Frail<br>(n=119) | P-value |
| **Age** | 77.9<br>(6.83) | 77.9<br>(6.64) | 77.9<br>(7.04) | 0.91 | 73.0<br>(5.86) | 73.2<br>(6.12) | 69.8<br>(4.61) | 0.63 |
| **Number of females** | 82<br>(39%) | 41<br>(39%) | 41<br>(39%) | 1.00 | 82<br>(34.4%) | 41<br>(34.4%) | 41<br>(34.4%) | 1.00 |
| **BMI** | 26.29<br>(6.28) | 26.06<br>(6.32) | 26.5<br>(6.28) | 0.82 | 26.8<br>(5.00) | 26.8<br>(5.09) | 26.8<br>(4.92) | 0.94 |
| **Cardiovascular disease** | 140<br>(66.0%) | 71<br>(67.0%) | 69<br>(65.0%) | 0.88 | 37<br>(15.5%) | 28<br>(23.5%) | 9<br>(7.6%) | 0.001 |
| **Diabetes** | 85<br>(40.4%) | 41<br>(39.0%) | 44<br>(42.0%) | 0.67 | 38<br>(16.0%) | 25<br>(21.0%) | 13<br>(10.9%) | 0.051 |
| **Respiratory disease** | 84<br>(40.0%) | 52<br>(49.0%) | 32<br>(30.0%) | 0.007 | 50<br>(21.0%) | 35<br>(29.4%) | 15<br>(12.6%) | 0.002 |
| **Psychiatric condition** | 23<br>(11.0%) | 15<br>(14.0%) | 8<br>(7.0%) | 0.18 | - | - | - | - |
| **Dementia** | 40<br>(19.0%) | 35<br>(33.0%) | 5<br>(4.7%) | 1.07x10<sup>-7</sup> | - | - | - | - |
| **Cancer<sup>a</sup>** | 39<br>(18.6%) | 19<br>(18.0%) | 20<br>(19.0%) | 0.86 | 27<br>(14.1%) | 25<br>(23.6%) | 2<br>(2.3%) | 1.61x10<sup>-7</sup> |

<sup>a</sup>Cancer data only available for 192 individuals (106 frail and 86 not frail) in the cohort.

**Figure 1:**
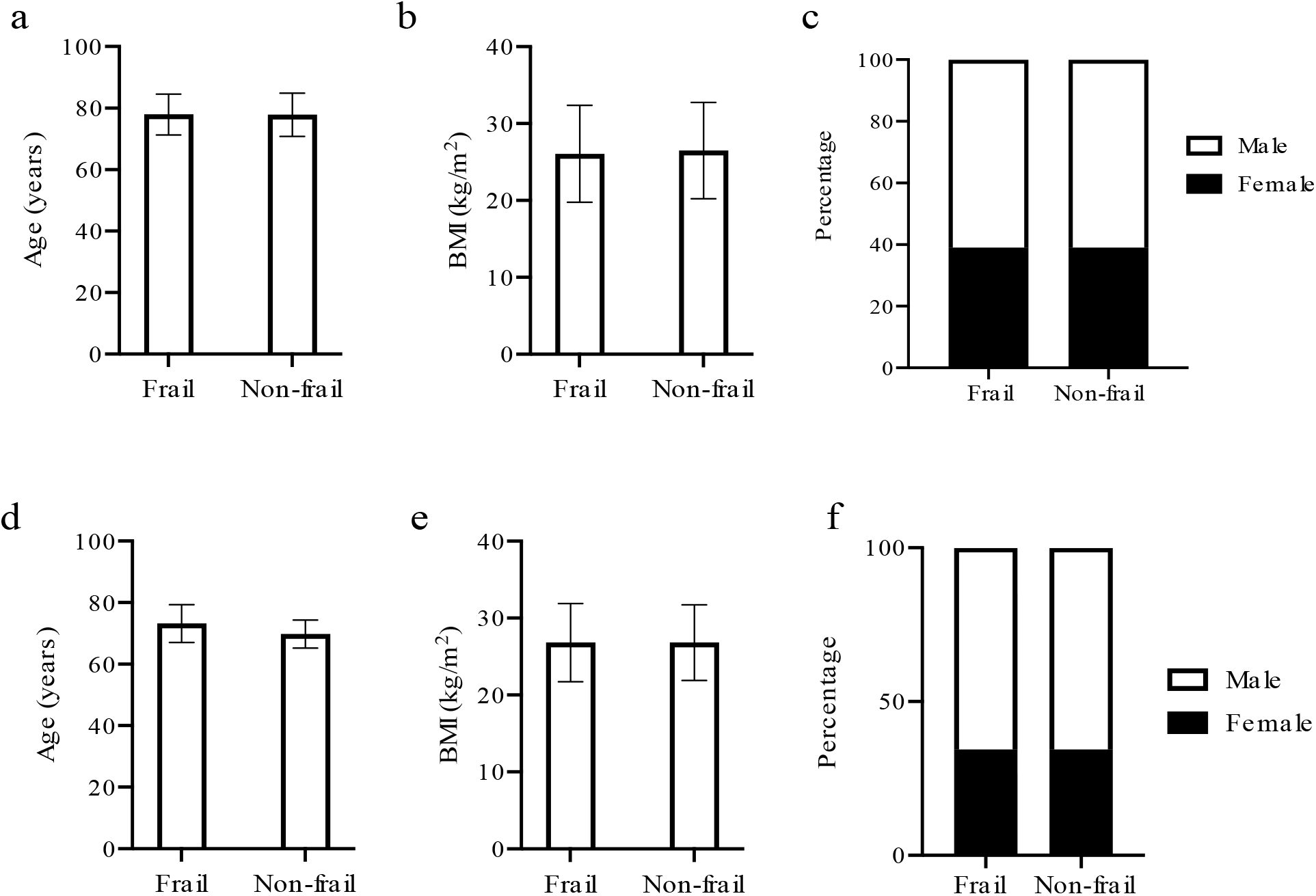
Baseline Characteristics of Study Population according to Frailty in Hospitalised and in Community-based Cohorts (Age Matched Populations) For all graphs: F = frail subgroup, NF = non-frail subgroup. (a) Mean age (in years), (b) mean BMI (kg/m2) F and NF groups in both sexes combined, female and males, (c) Percentage of females in F and NF groups in hospital cohort. (d) Mean age (in years), (e) mean BMI (kg/m2) F and NF groups in both sexes combined, female and males, (f) Percentage of females in F and NF groups in Community-based cohort.

After age-matching, delirium was reported in 40 (38%) of frail and 13 (12%) of non-frail patients with COVID-19 (Table 3). Frailty was found to significantly predict delirium (*P*-value: 0.013; Odds Ratio (OR) (95% Confidence Interval (CI)) = 3.22 (1.44, 7.21)) after false discovery rate (FDR) correction for multiple testing (Figure 2 and Table 3). There were no significant differences between frail and not frail for other symptoms (fever (temperature ≥ 37.5C) and cough).

**Table 3:**
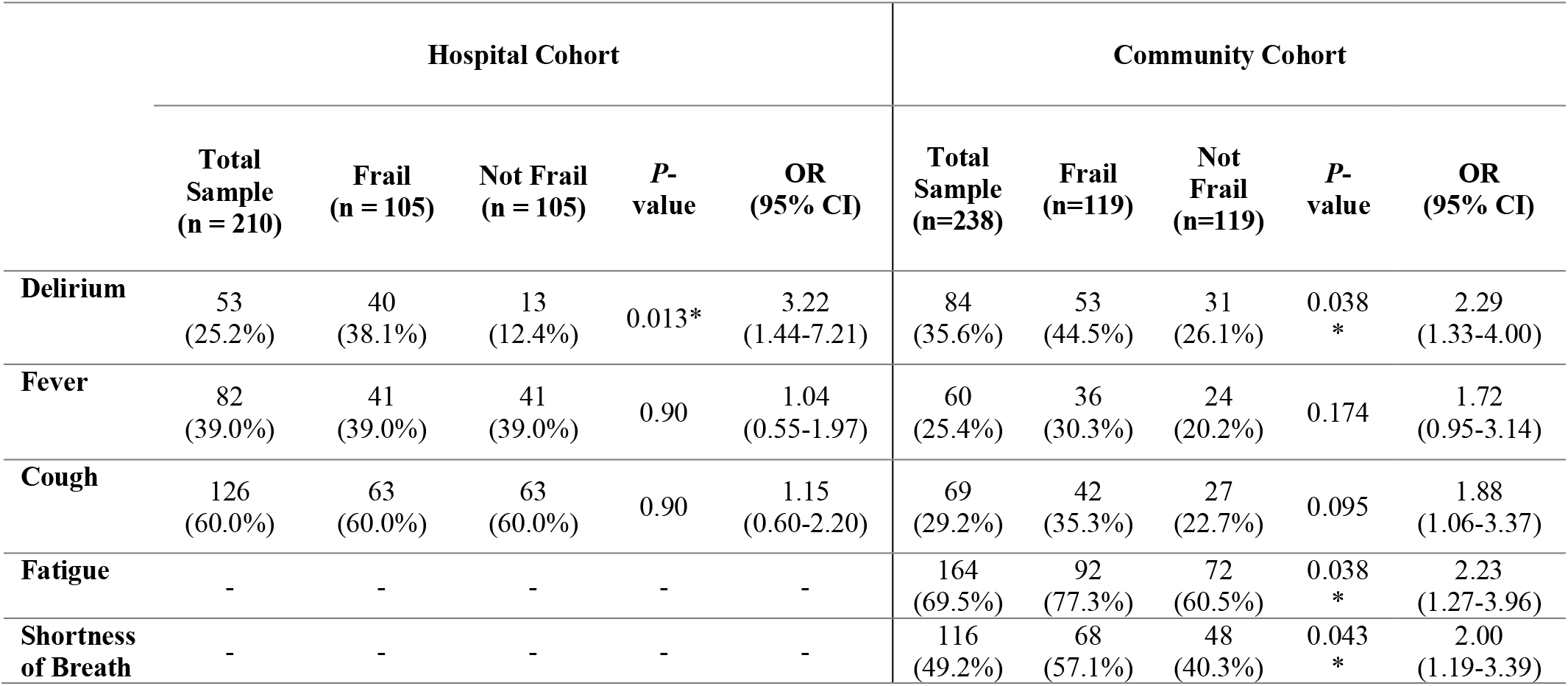
COVID-19 symptoms of age-matched hospital and community-based cohorts, according to frailty status. Binary variables are presented as count (%) and continuous variables as mean (standard deviation). Frail hospital = CFS ≥ 5; Frail community-based= PRISMA7 ≥3; Fever in hospital data (≥37.5°C) *P*-values are FDR adjusted at 5%. *donates significance

**Figure 2:**
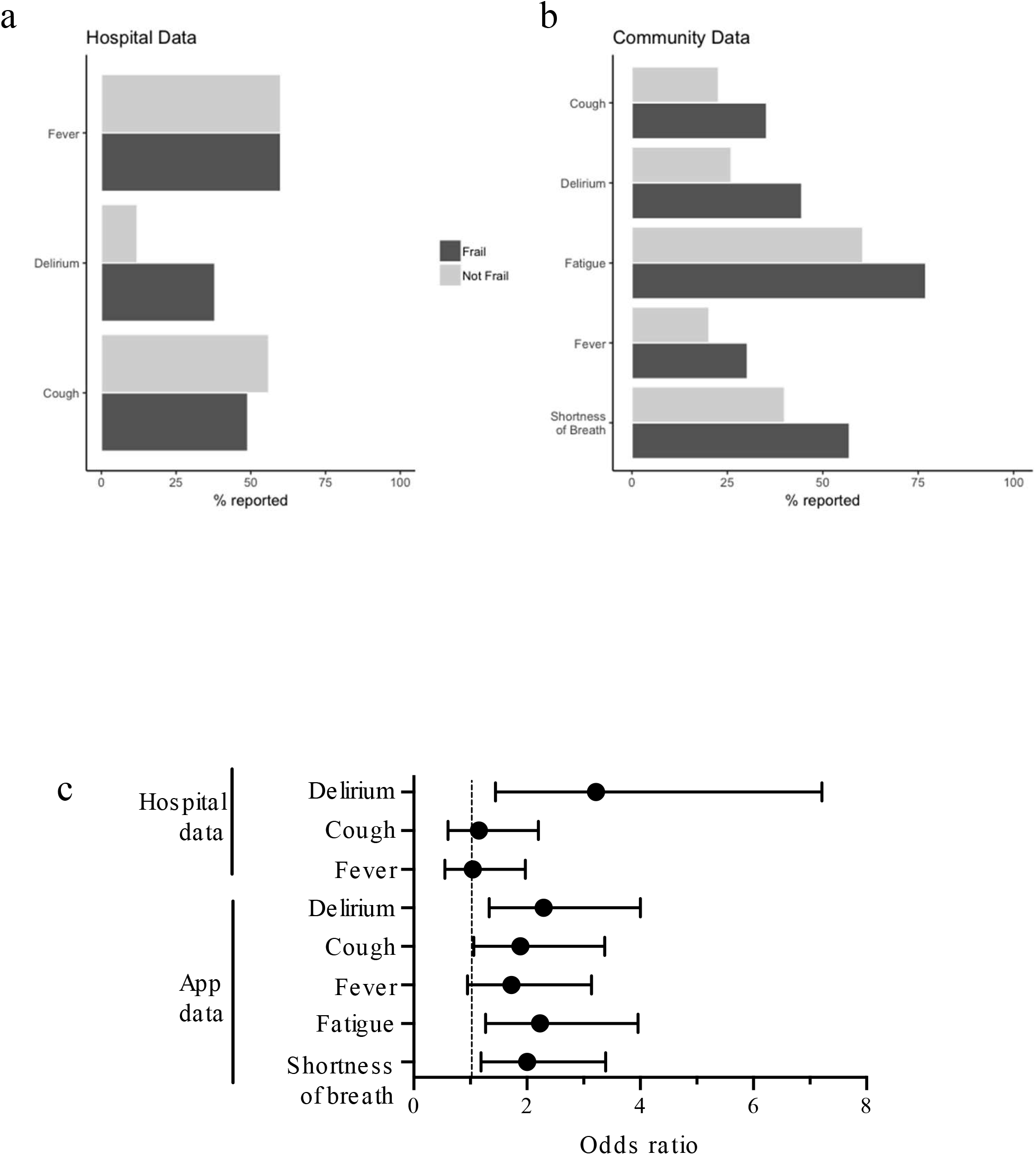
Proportions of frail and non-frail individuals presenting with symptoms in A) hospital cohort; B) community-based cohort, and C) Odds ratios of effect of frailty on presentation of symptoms.

### Community-based cohort

The proportion of total COVID Symptom Study app users ≥65 years of age was 334,544/ 2,848,396 (11.7%). Baseline characteristics of the unmatched study population are summarised in Supplementary Table SP2. 192 (36%) were classified as frail. 77% of the 192 frail participants with a reported positive COVID-19 test were reporting-by-proxy. In the frail group, delirium was present in 94/192 (49%) (self-reported 20/44 (45%); reported-by-proxy 74/148 (50%)). Baseline characteristics of the age-matched cohort are summarised in Figure 1 (d-f) and Table 2. Comorbidities are summarised in Table 2.

After age-matching, frailty was found to significantly predict delirium (*P*-value 0.038; OR (95% CI) = 2.29 (1.33, 4.00)), after FDR correction for multiple testing (Table 3). Frailty also predicted fatigue (*P*-value: 0.038; OR = 2.23 (1.27, 3.96)) and shortness of breath (*P*-value: 0.043; OR = 2.00 (1.19, 3.39)). There were no significant differences between frail and not frail for the other 11 symptoms analysed (Table 3 and Supplementary Table SP3).

## Discussion

In this study, we demonstrate that prevalence of delirium and confusion is significantly higher in frail compared to non-frail older adults with COVID-19, highlighting both that a frailty assessment is fundamental, but also a systematic evaluation of change in mental status needs to be included when assessing this population. Delirium is associated with greater morbidity and mortality amongst older adults[27] and may be the only presenting symptom of COVID-19 in this highly vulnerable population.

COVID-19 disproportionately affects the older population, partly because of the increased prevalence of frailty. Frail adults in residential settings may be at particular risk of transmission of respiratory illness. Case reports of COVID-19 outbreaks in nursing and residential homes have been published, highlighting the risks of infection in residential settings and a high case fatality[28]. Aside from residential facilities, many frail older adults are dependent on home care packages, with movement of care workers between residences. It is imperative that differences in COVID-19 presentation as a result of frailty are understood to prompt rapid diagnosis, isolation and contact tracing to limit outbreaks, and to clinical assessment and treatment. Our findings alert clinicians to the need to identify frailty and look for atypical COVID-19 presentations.

We also identified self-reported fatigue and shortness of breath as more common amongst adults with frailty. Higher prevalence of fatigue is consistent with a decrease in physiological reserve conferred by frailty.

Early identification of frailty facilitates a differential, targeted clinical approach, focusing not only on “typical” COVID-19 symptoms (*e*.*g*. fever and cough) but also on symptoms, such as delirium, demonstrated here to be more common in frail older adults. The importance of comprehensive frailty assessment in acute clinical settings and the community is nationally recognised[29,30]. Rapid NICE guidance produced in response to the COVID-19 outbreak outlines the importance of identifying those at increased risk of poor outcomes and perhaps less likely to benefit from critical care admission[18,30]. Delirium is frequently under-recognised in hospitalised populations[27]. Recognition of delirium necessitates a thorough investigation for underlying causes and management requires a tailored, patient-centred approach.

Delirium is a frequent complication of hospitalisation for older adults[31] and for patients admitted to critical care of all ages[32]. Whilst pathophysiology is not fully understood, neuroinflammation has been hypothesised to play a key role in its development[32]. In the context of COVID-19, few of the early descriptive studies included delirium or new confusion[33], emphasising the importance of our finding. A recent case study described acute confusion in an older patient with no pre-existing cognitive impairment presenting with COVID-19[14]. In a retrospective study of 214 patients from China, Mao *et al*. described neurological manifestations of COVID-19 and reported ‘impaired consciousness’ in 7.5% of patients, some of which may represent delirium. Other central nervous system (CNS) manifestations of COVID-19 infection reported were headache, dizziness, acute cerebrovascular disease, ataxia and seizures. These were significantly more common in severe infections and in older and comorbid patients[34]. In addition, non-neurological triggers may be important in delirium. Hypoxia, common in COVID-19 infections[35], is a well-recognised cause of delirium[36]. In addition, electrolyte disturbance, dehydration, and tachycardia common to all viral infections may play a role. Some have suggested that reduced patient contact during the pandemic, for protection of staff and other patients and because of time taken to don personal protective equipment, may be contributing to missed or delayed diagnoses of fluctuant delirium and acute confusion in these patients[37,38].

A number of other coronaviruses have shown significant neurotropism in animal models [39,40]; it is plausible that SARS-CoV-2 displays this property. Indeed, SARS-CoV-2 was found in cerebrospinal fluid of a reported case of COVID-19 encephalitis in Beijing[41]. Different mechanisms have been proposed to explain neurological involvement in COVID-19. The virus could reach the CNS via a compromised blood-brain barrier, or migrate via neuronal pathways, infecting peripheral sensory or motor nerve endings[42], leading to subsequent neurological damage and neuroinflammation. Some authors have proposed that angiotensin-converting enzyme 2 (ACE2), identified as the functional receptor for SARS-CoV-2, also expressed in the brain[43], could play a crucial role in mediating the inflammatory response in COVID-19 infection[34,44].

### Limitations

There are several limitations to this study. For the hospital cohort, admissions data were collected retrospectively from electronic health records. CFS was not always documented systematically and, in approximately one-third of cases, had to be retrospectively calculated using relevant information from admission clerking.

In the community-based cohort, COVID-19 test result and symptoms were self-reported and entered directly by participants or by-proxy. The proportion of total app users ≥65 years of age is 11.7%, whereas only 4.8% of app users with a confirmed positive COVID-19 test were ≥65 years of age. Since 18.3% of the UK population are aged over 65[45], older adults overall, and especially laboratory-tested for COVID-19, are likely to be under-represented. Sampling using a mobile application will under-represent individuals without mobile devices and is likely to under-represent those severely affected by the disease. These biases may mean we underestimate the differences in symptoms in frail people in the community study. The fact that the most significant finding from the community-based study was recapitulated in the clinical study is reassuring. Furthermore, of app users ≥65 years of age, 16.4% are categorised as frail. This is a similar proportion to the 15-20% of older adults categorised as moderate or severely frail during validation of the electronic Frailty Index[46].

For community-based data, note that COVID-19 tests were not widely available in the UK at the time of study[47,48]. Participants were likely tested because they displayed severe symptoms, were in contact with confirmed cases, were healthcare workers or had travelled abroad to a risk area early in the outbreak. Additionally, the app captures data on whether participants have ever been diagnosed with COVID-19; some individuals may have previously tested positive but were no longer symptomatic when they enrolled.

## Conclusion

This is the first study reporting that frail patients with COVID-19 present with a significantly higher prevalence of delirium than non-frail patients, using hospital and community-based cohorts. Our data emphasises the need for systematic assessment of frailty and delirium as critical components when assessing acutely ill patients in both hospital and community settings. Clinicians should suspect COVID-19 in frail patients presenting with delirium. This is especially important in care homes and long-term care facilities, where resident populations are vulnerable to respiratory disease outbreaks. Early detection facilitates implementation of infection control measures and appropriate personal protective equipment to avoid catastrophic spread and reduce mortality amongst frail older adults.

## Ethics

COVIDCollab project service evaluation approved by Guy’s and St Thomas’ NHS Trust audit leads. Reference number 10777.

The Covid Symptom Study App Ethics has been approved by KCL ethics Committee REMAS ID 18210, review reference LRS-19/20-18210. All subscribers provided consent when signing up for the app.

## Data sharing

COVIDCollab data is available upon application to the Geriatric Medicine Research Collaborative https://www.gemresearchuk.com/.

App data used in this study is available to bona fide researchers through UK Health Data Research using the following link https://healthdatagateway.org/detail/9b604483-9cdc-41b2-b82c-14ee3dd705f6

## Data Availability

COVIDCollab data is available upon application to the Geriatric Medicine Research Collaborative https://www.gemresearchuk.com/.
App data used in this study is available to bona fide researchers through UK Health Data Research using the following link https://healthdatagateway.org/detail/9b604483-9cdc-41b2-b82c-14ee3dd705f6

https://healthdatagateway.org/detail/9b604483-9cdc-41b2-b82c-14ee3dd705f6

## Acknowledgements

The app was developed by Zoe Global Limited with input from King’s College London and Massachusetts General Hospital.

The Principal Investigator and corresponding author (MNL) had full access to all the data in this study and takes full responsibility for the integrity of the data and the accuracy of data analysis.

All authors listed were responsible for aspects of study design, data collection and analysis and writing of manuscript and meet criteria for authorship. MBZ, RSP, ALR, CW, HD and MNL were involved in data collection, cleaning and statistical analysis of the hospital dataset. BM produced the data cleaning script for the app dataset. ALR, KAL, CHS, RCEB, AV, MM, MBF, JSESM, KS, MM, JW and SO were involved in data collection, processing and analysis of the app dataset. MBZ, RSP, ALR, KAL, HD, CHS, CW, RCEB, AV, MM, FCM, CJS and MNL drafted the initial manuscript. All authors contributed to and reviewed the final manuscript to be published.

We would like to acknowledge the following COVIDCollab collaborators, responsible for hospital data collection: Rishi Iyer, Rachael Anders, Lindsay Hennah, Gitanjali Amaratunga, Abigail Hobill, Cassandra Fairhead, Amybel Taylor, Henry Maynard, Marc Osterdahl, Maria Dias, Taha Amir, Natalie Yeo, Jamie Mawhinney, Hamilton Morrin, Li Kok, Luca Scott, Aiden Haslam, Gavriella Levinson, Stephanie Mulhern, Stephanie Worrall, Thurkka Rajeswaran, Katherine Stamboullouian, Sophie McLachlan, Karla Griffith, Daniel Muller, Alice O’ Doherty, Baguiasri Mandane, Irem Islek, Alexander Emery, John Millwood-Hargrave, Andra Caracostea, Laura Bremner, Arjun Desai, Aneliya Kuzeva, Carolyn Akladious, Mettha Wimalasundera, Mairead Kelly, Sally Aziz, Sinead O’Dwyer, Rupini Perinpanathan, Anna Barnard, Nicole Hrouda, Ismini Panayotidis, Nirali Desai, Hannah Gerretson, Rebecca Lau, Zaynub Ghufoor, Hanna Nguyen, Torben Heinsohn, Jack Cullen, Eleanor Watkins, Vaishali Vyas, Daniel Curley, Niamh Cunningham, Vittoria Vergani, Kelvin Miu, Jack Stewart, Nicola Kelly, Lara Howells, Benyamin Deldar, Ross Sayers, Gracie Fisk, Sri Sivarajan, Tahmina Razzak, Helen Ye, Samiullah Dost, Nikhita Dattani, Catherine Wilcock, Gabriel Lee, Jodie Acott, Hannah Bridgwater, Antia Fernandez, Hesham Khalid, Katherine Hopkinson, Deirdre Green, Hejab Butt, Ayushi Gupta, Madeleine Garner, Hazel Sanghvi, Madeleine Daly, Emily Ross-Skinner, Shefali Patel, Danielle Lis. (All affiliated to Guy’s and St Thomas’ Hospital NHS Foundation Trust).

Written permission obtained to include the names of all individuals listed.

The authors acknowledge use of the research computing facility at King’s College London, *Rosalind* (https://rosalind.kcl.ac.uk).

